# Prior Optic Neuritis Detection on Peripapillary Ring Scans using Deep Learning

**DOI:** 10.1101/2022.04.27.22274388

**Authors:** Seyedamirhosein Motamedi, Sunil Kumar Yadav, Rachel C. Kenney, Ting-Yi Lin, Josef Kauer-Bonin, Hanna G. Zimmermann, Steven L. Galetta, Laura J. Balcer, Friedemann Paul, Alexander U. Brandt

## Abstract

**Background:** The diagnosis of multiple sclerosis (MS) d requires demyelinating events that are disseminated in time and space. Peripapillary retinal nerve fiber layer (pRNFL) thickness as measured by optical coherence tomography (OCT) distinguishes eyes with a prior history of acute optic neuritis (ON) and may provide evidence to support a demyelinating attack.

**Objective:** To investigate whether a deep learning (DL)-based network can distinguish between eyes with prior ON and healthy control (HC) eyes using peripapillary ring scans.

**Methods:** We included 1,033 OCT scans from 415 healthy eyes (213 HC subjects) and 510 peripapillary ring scans from 164 eyes with prior acute ON (140 patients with MS). Data were split into 70% training (728 HC and 352 ON), 15% validation (152 HC and 79 ON), and 15% test data (153 HC and 79 ON). We included 102 OCT scans from 80 healthy eyes (40 HC) and 61 scans from 40 ON eyes (31 MS patients) from an independent second center. Receiver operating characteristic curve (ROC) analyses with area under the curve (AUC) were used to investigate performance.

**Results:** We used a dilated residual convolutional neural network with alternating convolutional and max pooling layers for the classification. A final network using 2-factor augmentation had an accuracy of 0.85. The network achieved an area under the curve (AUC) of 0.86, whereas pRNFL only had an AUC of 0.77 in recognizing ON eyes. Using data from a second center, the network achieved an accuracy of 0.77 and an AUC of 0.90 compared to pRNFL, which had an AUC of 0.84.

**Conclusion:** DL-based disease classification of prior ON is feasible and has the potential to outperform thickness-based classification of eyes with and without history of prior ON.

## Introduction

Multiple sclerosis (MS) is the most common chronic autoimmune demyelinating disorder of the central nervous system^1^. In the absence of a pathological biomarker, diagnosing clinically definite MS (CDMS) requires dissemination of demyelination and neurological symptoms in time and space^2^. The McDonald criteria allow the diagnosis of MS at the time of a clinically isolated syndrome (CIS) by utilizing biomarkers that primarily include T2-weighted lesions on brain magnetic resonance imaging (MRI) to establish paraclinical evidence of dissemination in time and space^3^. While the McDonald criteria are ubiquitously used in clinical care, their sensitivity and specificity for clinically definite MS are only moderate^4^ causing delays in diagnosis^5^ and diagnosticcertainty^6^. Further, longitudinal studies have shown earlier treatment of ON with MS-disease modifying therapies improves visual and physical disability over time, rendering earlier diagnosis of MS by adding ON as a lesion site important^7^.

The optic nerve is a key site of disease in MS^8^. Post-mortem studies reveal significant retinal ganglion cell loss in more than 70% of MS patient eyes^9,10^. While acute optic neuritis (ON) represents the initial manifestation of MS in up to 25% of patients, and approximately 50% of MS patients experience acute ON at some point in their disease, demyelinating plaques in the optic nerve are found in up to 99% of MS patients at *post mortem*^11,12^. Recognizing optic nerve pathology, i.e. signs of prior ON, could thus serve as an additional criterion supplementing the current McDonald criteria^13^. During the last revision of the criteria this potential was recognized, but optic nerve assessment was ultimately not included in the revised criteria because of a lack of supporting data, which were mainly derived from MR imaging studies that may or may not have included dedicated orbital images or of the optic nerve with fat-saturation and contrast^14^.

Optical coherence tomography (OCT) obtains high-resolution structural measurements of retinal thickness that reflect the integrity of the optic nerve. These measurements have been suggested as diagnostic biomarkers in MS that not only reflect structural but also functional visual outcomes in MS^15–17^. As such, the peripapillary retinal nerve fiber layer thickness (pRNFL) is reduced within weeks to months after ON, and increased magnitudes of thickness reduction are associated with worsening degrees of visual function loss^18^. However, the large individual ranges of normal OCT measurements render absolute thickness measurements of pRNFL alone less useful for detecting prior ON^19,20^.

Artificial intelligence, using deep learning and convolutional neural networks, allow image classification based on raw imaging data^21^. Deep learning-based methods are trained directly on imaging data, and resulting networks have shown remarkable success in recognizing retinal pathologies in primary eye disorders^22^. Recently, we have shown that deep learning can be used for automatic quality control of OCT images^23^. We and others have further shown that deep learning-based image segmentation outperforms classical intra-retinal layer segmentation on OCT images^24^.

We hypothesized in this study that deep learning-based recognition of prior ON by peripapillary ring scans is feasible and outperforms thickness-based classification using absolute pRNFL values. To investigate this, we trained a deep neural network on a carefully-curated data set of pRNFL scans from healthy subjects and eyes with history of acute ON of patients with MS. We then evaluated network performance for recognizing prior ON under several conditions, including independent validation data.

## Methods

### Study population

In this study, we retrospectively included data from relapsing-remitting MS (RRMS) and CIS patients with a history of ON from two longitudinal observational studies at the Experimental and Clinical Research Center (ECRC) of Charité - Universitätsmedizin Berlin. Data were acquired between April 2011 and September 2021. Inclusion criteria were minimum age of 18 years and a diagnosis of RRMS or CIS according to the 2017 revised McDonald criteria^3^. Exclusion criteria were acute ON within 90 days before OCT examinations and any other neurological or eye disorder known to affect the retina (e.g. glaucoma, diabetes, and refractive error of greater than 6 diopters). We only included eyes with a documented history of ON; this was based on providers’ notes in medical records and patient attack history. Additionally, age- and sex-matched data from healthy controls (HC) with no history of neurological or eye disorder from the ECRC’s database were included in this study. Repeated measurements during follow-up were allowed in this study. Table 1 shows an overview of the study cohort.

**Table 1:** Demographic description of HC and ON cohorts. Abbreviations: HC = Healthy controls; ON = Optic neuritis; OCT = Optical coherence tomography; RRMS = Relapsing-remitting multiple sclerosis; CIS = Clinically isolated syndrome. a) Age match: p value = 0.12, b) Sex match: p value = 1

|  | HC | ON |
| --- | --- | --- |
| No. of subjects [N] | 213 | 140 |
| No. of eyes [N] | 415 | 164 |
| No. of OCT Scans [N] | 1033 | 510 |
| Age at baseline [years]<sup>a</sup> | 36 ± 12 | 37 ± 10 |
| Sex (female) [N (%)]<sup>b</sup> | 142 (67%) | 93 (66%) |
| Time since onset at baseline (median [range]) [years] | -- | 1.12 [0.26 – 43.08] |
| Time since last ON (median [range]) [years] | -- | 3.02 [0.25 – 44.83] |
| Diagnosis at baseline | -- | 108 RRMS 32 CIS |

A confirmatory cohort, consisting of 61 OCT scans from 40 eyes of 31 patients with RRMS with a history of ON (65% women, age at baseline: 41±12 years) and 102 OCT scans from 80 eyes of 40 healthy controls (53% women, age at baseline: 29±10 years) from a longitudinal observational cohort study at the Department of Neurology, New York University Grossman School of Medicine, New York, NY, was included in this study by incorporating the same inclusion and exclusion criteria. RRMS and HC groups were well matched in the confirmatory cohort by sex (*p* value: 0.43) but not by age (*p* value: <0.001). The mean time from disease onset for the RRMS group was 10.02 years with minimum of 0.25 and maximum of 23 years.

### Optical coherence tomography

All OCT scans of both exploratory and confirmatory cohorts were performed using Spectralis spectral-domain OCT (Heidelberg Engineering, Heidelberg, Germany), with automatic real-time (ART) averaging and activated eye tracking. Measurements were performed in a dimly lit room without pupil dilation unless required. Peripapillary ring scans were obtained either as single 2D OCT B scans (12º, 1536 A scans per B scan, 496 pixels per A scan, 16 ≤ ART ≤ 100) or extracted from the optic nerve head radial and circular (ONH_RC) protocol taken with anatomic positioning system (APS) (inner ring scan with constant circle diameter of 3.5 mm, 768 A scans per B scan, 496 pixels per A scan, 16 ≤ ART ≤ 100). All scans were quality controlled according to the OSCAR-IB criteria^25^ by an experienced grader. Seven OCT scans from the exploratory cohort and one OCT scan from the confirmatory cohort were excluded from the study because of inadequate OCT scan quality or inaccurate intraretinal segmentation. OCT data are reported in accordance with the Advised Protocol for OCT Study Terminology and Elements (APOSTEL) recommendations^26^. This study was conducted in accordance to the artificial intelligence extension of the OSCAR-IB criteria (OSCAR-AI)^27^, with the exception that the criterion of Openness could not be fulfilled because of data protection constraints on data-sharing.

### Artificial neural network

#### Training test and validation data sets

The OCT scans included in this study were randomly allocated to the training, validation and test data sets. We allocated 70% of the data for model training, 15% for model validation during the training and 15% for model testing after the training. Data allocation of repeated measures of an individual subject required that all measurements of this individual only appeared in one data set. Random allocation was repeated until having around 70% of patients and eyes in the training data set and 15% of them in each of the validation and test data sets. This process was necessary in order to ensure the homogeneity of all data sets with regard to the number of patients and eyes in addition to the number of OCT scans. In the end, we allocated 728 OCT scans from 290 eyes of 149 HC and 352 OCT scans from 115 eyes of 98 patients to the training data set, 152 OCT scans from 32 HC (63 eyes) and 79 OCT scans from 21 patients (25 eyes) to the validation data set, and 153 OCT scans from 32 HC (62 eyes) and 79 OCT scans from 21 patients (24 eyes) to the test data set.

#### Preprocessing

Figure 1 summarizes all preprocessing steps. All OCT scans went through intraretinal segmentation of inner limiting membrane (ILM), lower boundary of retinal nerve fiber layer, and Bruch’s membrane (BM) using Nocturne ONE software (Nocturne GmbH, Berlin, Germany). All segmentation results were manually quality controlled as described above. Based on the segmentation, a label image was created for each OCT scan with four different classes: 1) the area above the ILM, 2) pRNFL, 3) the area between the lower boundary of RNFL and BM, and 4) the area below BM. After segmentation, both OCT and label images were flattened using the BM as reference. Scans were then resized to a fixed size of 512×512 pixel using bi-linear interpolation. Finally, the intensity of pixels located between ILM and BM (foreground) was linearly normalized for each individual scan, and the intensity of the pixels above ILM and below BM (background) was set to zero (black background).

**Figure 1:**
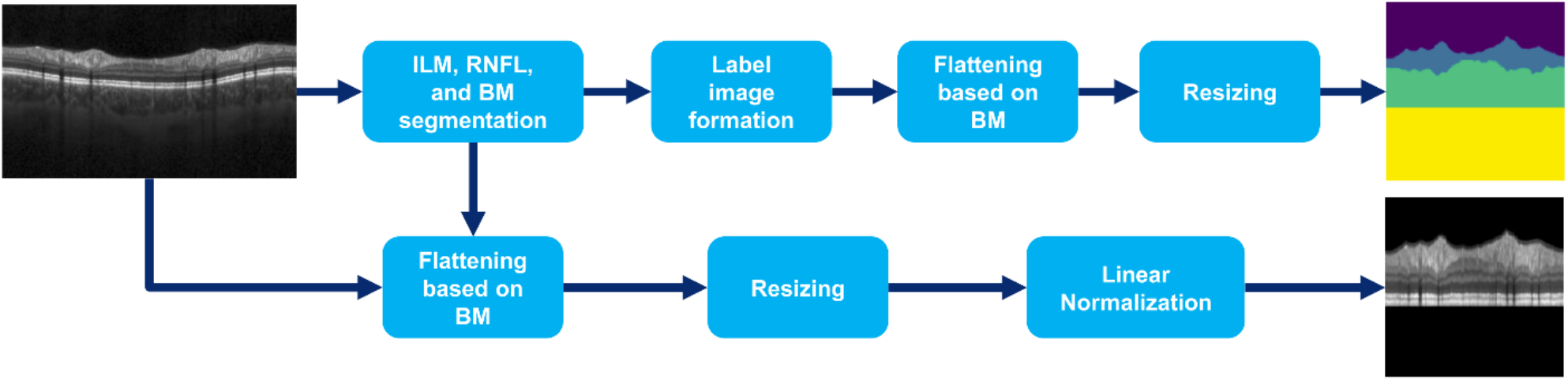
The pipeline for the pre-processing step. Abbreviations: ILM = Inner limiting membrane; RNFL = Retinal nerve fiber layer; BM = Bruch’s membrane.

#### Data augmentation

To increase data size, diversity, translation invariance, classification accuracy and possibly reduce over-fitting, we performed model training additionally on synthetically modified data created using data augmentation. Augmentation was performed by applying random horizontal and vertical circular shift of up to 20 pixels to all OCT images of the training data set. To test if data augmentation results in improvements, the model was trained on five different training data sets with these scenarios: 1) no data augmentation (510 ON OCT images, 1033 HC OCT images),

2) ON data augmentation by a factor of 2 using random vertical shift (1020 ON OCT images, 1033 HC OCT images), 3) ON data augmentation by a factor of 2 using random vertical and horizontal shift (1020 ON OCT images, 1033 HC OCT images), 4) ON data augmentation by a factor of 4 and HC data augmentation by a factor of 2 using random vertical shift (2040 ON OCT images, 2066 HC OCT images), and 5) ON data augmentation by a factor of 4 and HC data augmentation by a factor of 2 using random vertical and horizontal shift (2040 ON OCT images, 2066 HC OCT images). The main reason to augment more on ON data was that repeated OCT measurements at follow ups are more similar to each other in healthy controls than ON patients who have a possibility of having disease worsening and repeated acute ON episodes between the measurements. Another reason behind performing more data augmentation on ON was to compensate for the imbalance between the size of the HC and ON data. The reason behind applying mainly vertical shift was that OCT ring scans have more positioning similarity in the horizontal direction than the vertical direction.

#### Implementation

The model was fully developed in Python version 3.7 using Keras with TensorFlow backend. Early stopping regularization was used to avoid over-fitting during training by stopping the training process after the convergence of training and validation accuracy. Model training was performed on an on-premise Linux workstation with two Intel Xeon Gold 6144 (Intel Corporation, Santa Clara, CA, USA) CPUs and four NVIDIA GeForce GTX 1080 Ti (NVIDIA Corporation, Santa Clara, CA, USA) GPUs. Model testing was performed on a Windows desktop with an Intel Core i9 11980HK CPU and a NVIDIA GeForce RTX 3080 GPU.

#### Statistical analysis

The convenience sample size in this pilot study was not based on an *a priori* sample size estimation. To confirm group matching, sex differences between groups were tested using χ^2^ test and age differences were tested using 2-sample Wilcoxon test. Classification performance was evaluated using area under the curve (AUC) from receiver operating characteristic (ROC) curves. The optimal cutoff for the classifier was estimated based on the intersection of a 45º tangent line and the ROC curve, which is equivalent to the maximum of the summation of the sensitivity and specificity. F_1_ score, a measure of accuracy, was calculated as: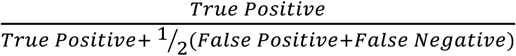, with ON as the positive class and HC as the negative. *p* values less than 0.05 were considered significant. All statistical analysis were performed in R version 3.5^28^.

#### Ethics statement

The study was approved by the local ethics committee at Charité — Universitätsmedizin Berlin (EA1/182/10 and EA1/163/12). Independent validation data from NYU were collected under IRB approval at NYU. The study was conducted according to the Declaration of Helsinki in its currently applicable version and the applicable German and European laws. All participants gave written informed consent.

## Results

### Network architecture

The proposed convolutional neural network (CNN) for the classification of eyes with a history of ON vs HC eyes is shown in Figure 2. The designed CNN accepts two-channel inputs consisting of the OCT scans (channel 1) as well as their segmentation map (channel 2), both with dimensions of 512×512 pixels. Our proposed model uses dilated convolution, which results in exponential expansion of the receptive field by linearly increasing the dilation factor, without increasing the kernel size^29^. The model also employs residual learning using skip connections, which helps it to avoid the vanishing gradients problem^30^. The network has five dilated residual blocks, each consists of two convolutional layers with kernel size of 3×3 and different dilation factors, followed by concatenation of the feature maps created by each convolutional layer, a max-pooling layer of size 2×2 and stride of 2, and a dropout layer with dropout rate of 0.4. After the dilated residual blocks, the model has another 3×3 dilated convolutional layer with a dilation factor of 32, followed by max-pooling and dropout layers with the same features as the previous layers. Each convolutional layer is followed by a rectified linear unit (ReLU) activation function. In order to make the final prediction, the output of the last layer is flattened and then fed to a dense layer, followed by a sigmoid activation function. For the training, binary cross entropy (BCE) loss was used as 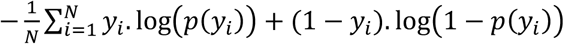 where *y* is the classification label (1 for ON and 0 for HC) and *p*(*y*) is the output of the model which is the predicted probability of an input image being from an eye with a history of ON. Adaptive moment estimation (Adam) algorithm was used to optimize the model during the training with a fixed learning rate of 0.0001.

**Figure 2:**
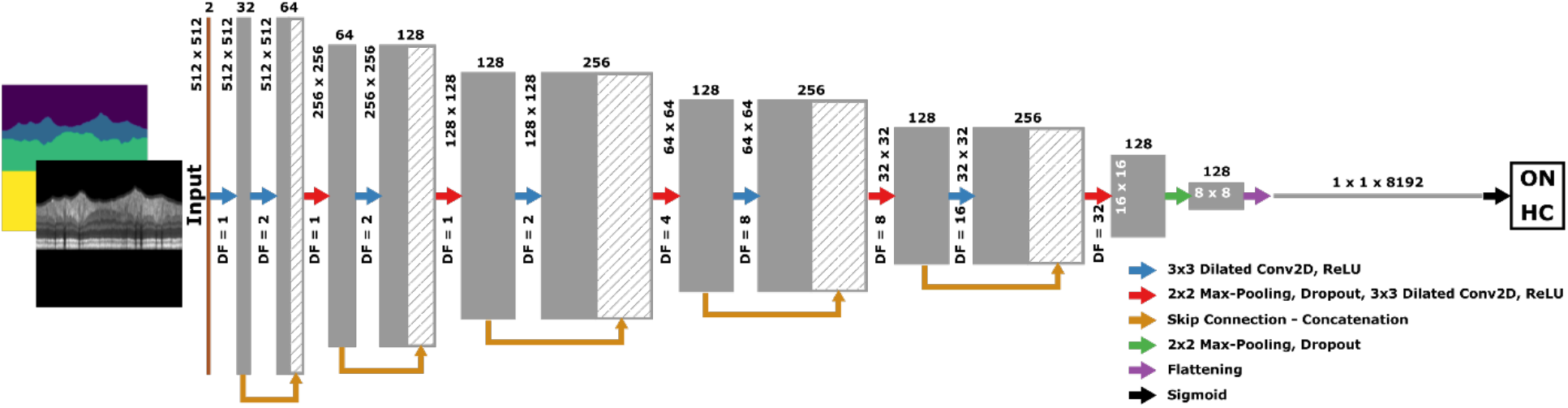
The architecture of the proposed model. Abbreviations: DF = Dilation factor; ON = Optic neuritis; HC = Healthy controls; Conv2D = 2D convolution layer; ReLU = Rectified linear activation function.

### Model validation with data augmentation

The proposed model was trained on training data sets with different data augmentation scenarios. Table 2 summarizes the performance of the model on the validation data set. As shown in the table, the models trained on the training data sets with only ON data augmentation have better overall performance in identification of eyes with prior ON from healthy controls, and therefore these models and the model trained on the training data set without any augmentation were selected to be tested on the test data set.

**Table 2:** Model performance with data augmentation on validation data set. Abbreviations: HC = Healthy controls - negative class; ON = Optic neuritis - positive class; Vert. = Vertical shift; Horiz. = Horizontal shift; TP = True positive; FN = False negative; FP = False positive; TN = True negative; Acc. = Accuracy; Sen. = Sensitivity; Spec. = Specificity; F1 = F1 score. The highest result for each evaluation metric is highlighted in bold.

| Aug. | Type | TP | FN | FP | TN | Acc. | Sen. | Spec. | F1 |
| --- | --- | --- | --- | --- | --- | --- | --- | --- | --- |
| None | -- | 44 | 35 | 6 | 146 | 0.82 | 0.56 | 0.96 | 0.68 |
| 2×ON | Vert. | 44 | 35 | 5 | 147 | **0.83** | 0.56 | 0.97 | **0.69** |
| 2×ON | Vert. -<br>Horiz. | 42 | 37 | 2 | 150 | **0.83** | 0.53 | **0.99** | 0.68 |
| 4×ON -<br>2×HC | Vert. | 45 | 34 | 11 | 141 | 0.81 | **0.57** | 0.93 | 0.67 |
| 4×ON -<br>2×HC | Vert. -<br>Horiz. | 43 | 36 | 5 | 147 | 0.82 | 0.54 | 0.97 | 0.68 |

### Recognizing signs of optic neuritis in MS eyes

Table 3 summarizes the performance of the selected models on the test data. The optimal cutoff for each classifier based on its performance on the test data was calculated and used to report the metrics in Table 3. As shown, the model trained on the training data set with only ON data augmentation with vertical shift had the best overall performance and thus was selected. The model was used with the classification cutoff calculated based on the test data in this study. ROC curve analysis for the selected classifier is shown in Figure 3a, next to ROC curve for pRNFL for comparison. Figure 4a shows pRNFL for both eyes with a history of ON and HC eyes, separated by the classification result.

**Table 3:** Model performance with data augmentation on test data set. Abbreviations: HC = Healthy controls - negative class; ON = Optic neuritis - positive class; Vert. = Vertical shift; Horiz. = Horizontal shift; Cut. = Optimal cutoff; TP = True positive; FN = False negative; FP = False positive; TN = True negative; Acc. = Accuracy; Sen. = Sensitivity; Spec. = Specificity; F1 = F1 score. The best result for each evaluation metric is highlighted in bold.

| Aug. | Type | Cut. | TP | FN | FP | TN | Acc. | Sen. | Spec. | F1 |
| --- | --- | --- | --- | --- | --- | --- | --- | --- | --- | --- |
| None | -- | 0.493 | 50 | 29 | 8 | 145 | 0.84 | 0.63 | **0.95** | 0.73 |
| 2×ON | Vert. | 0.469 | 58 | 21 | 14 | 139 | **0.85** | **0.73** | 0.91 | **0.77** |
| 2×ON | Vert. -<br>Horiz. | 0.467 | 53 | 26 | 9 | 144 | **0.85** | 0.67 | 0.94 | 0.75 |

**Figure 3:**
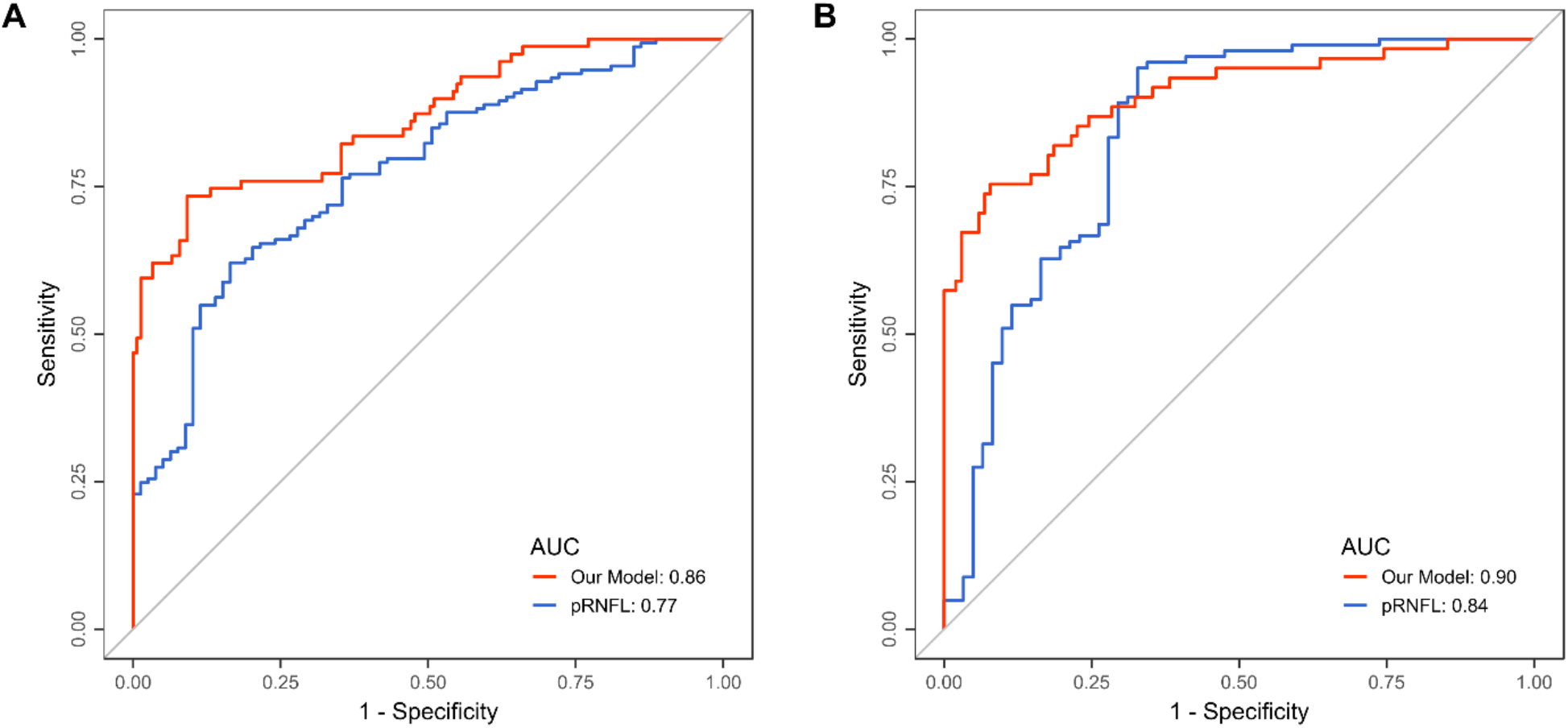
ROC curves for the proposed model and pRNFL identifying eyes with prior ON and HC in a) exploratory and b) confirmatory data. Abbreviations: ROC = receiver operating characteristic; AUC = Area under ROC curve; pRNFL = Peripapillary retinal nerve fiber layer thickness; ON = Optic neuritis; HC = Healthy control.

**Figure 4:**
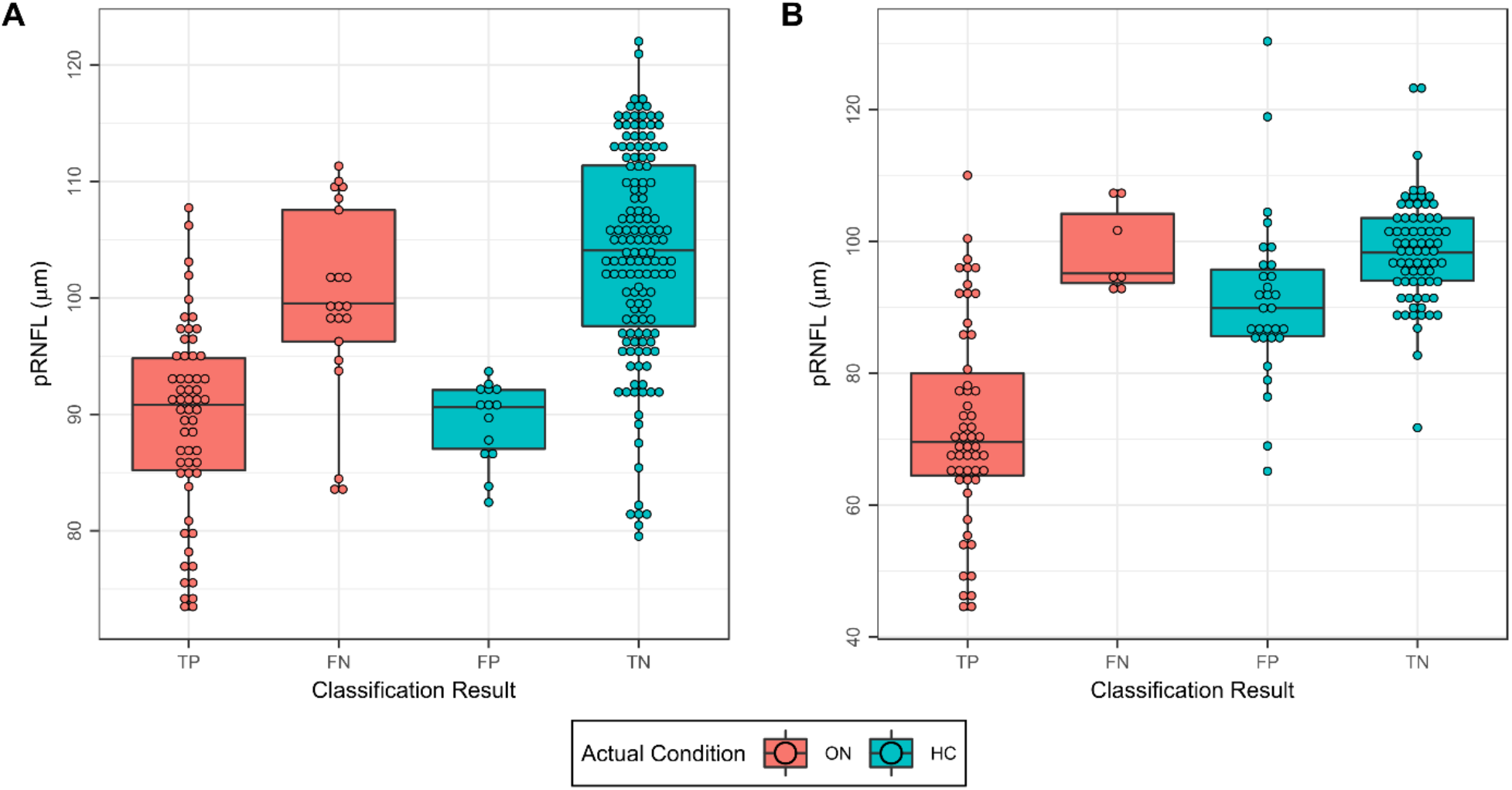
pRNFL stratified by classification result for a) exploratory and b) confirmatory data. Abbreviations: pRNFL = Peripapillary retinal nerve fiber layer thickness; ON = Optic neuritis; HC = Healthy control; TP = True positive; FN = False negative; FP = False positive; TN = True negative.

To account for correlation between repeated measurements per eye, the performance of the model was tested on a data set with only one OCT image per eye, randomly selected from the test data set. The model had an accuracy of 0.83 with the sensitivity of 0.67, specificity of 0.89, and F1 score of 0.68 for the classification of eyes with a history of ON and HC on the sub data set of the test data.

### Model confirmation

We tested our proposed model on an independent cohort of MS patients with a history of ON and HC, to investigate whether the results from the exploratory data could be confirmed on independent data. Our model had an accuracy of 0.77 with sensitivity of 0.89, specificity of 0.71, and F1 score of 0.74 (true positive: 54, false negative: 7, false positive: 30, true negative: 72). ROC curve analysis of the model on the confirmatory cohort as well as using pRNFL thickness to discriminate between ON and HC is shown in Figure 3b. Figure 4b shows the pRNFL values for this cohort against the classification result.

Similar to the exploratory data, we tested the performance of our model on a subset of the confirmatory cohort with only one OCT scan per eye, to account for the effect of repeated measurements per eye. Here, the model had an accuracy of 0.72 with sensitivity of 0.85, specificity of 0.65, and F1 score of 0.67.

## Discussion

In this study we show that deep-learning based classification of prior ON on peripapillary ring scans is feasible and outperforms pRNFL thickness-based classification in our initial cohort and a second independent data set. The good model performance in an independent cohort confirms the generalizability of our approach.

OCT is a simple, fast, and readily available method to query retina and optic nerve for neuroaxonal damage. OCT-derived measures, including pRNFL, have been suggested as imaging biomarkers for MS diagnosis^13^. Signs of prior ON have been estimated using inter-ocular differences (IOD) of retinal layers on the assumption that a typically unilateral ON would lead to neuroaxonal damage in one eye but not the other^31,32^. Percentage rather than absolute IOD may be superior over absolute IOD in detecting subtle changes^33^. Clinically meaningful cut-off values may be defined in comparison to healthy controls^20^ or based on large reference studies with MS-derived OCT measures^32,34^. Machine learning based classification of eyes with ON history utilizing standard OCT parameters has been promising (Kenney et al. *in press*). However, not all ON lead to measurable neuroaxonal damage, and in a previous study about 15% of patients do not suffer relevant damage during an ON episode^35^. This limits the value of methods relying on absolute OCT parameter thickness or inter-eye differences. Indeed, the proposed method recognized scans with normal pRNFL thickness correctly as ON, suggesting the DL-based classification may rely on other differences than only average thickness.

Other methods may provide alternative or complementary information regarding optic nerve pathology in MS. A conversion equation recently developed on an international cohort can provide a useful way to convert between two different OCT devices clinically and in research studies (Kenney et al. in press). Alternatively, i.e. optic nerve MRI^36^ or visual evoked potentials (VEP)^37^ may provide valuable biomarkers to determine dissemination in time and space criteria and for diagnosing MS.

Our choice for the convolutional neural network was a dilated residual network. Residual networks enhances the overall training with reduced training loss by applying residual skip connections to mainly avoid vanishing gradient problems^30^. Convolutional networks reduce the receptive field in deeper layers and make decisions based on small limited feature maps, leading to the loss of spatial information^29^. This problem can be avoided by either increasing the kernel size of convolutional layers or using the concept of dilated convolution. The latter option is preferable because it does not add more model parameters while increasing the receptive field, which is essential in our classification task, as the network needs to preserve spatial resolution to capture the structural relationship between different anatomical regions on peripapillary ring scans.

Our pilot study has notable weaknesses. Our network was trained on data from one device in one clinical center, whereas we were able to confirm model performance using additional data from a second, independent center. Thus, our study serves as a proof-of-principle, but the approach would need to be developed further to including multi-center, multi-device, multi-national and multi-racial/ethnic data to be clinically applicable. Comorbid diseases may affect retinal OCT scans and thickness measurements, and should also be considered^34^, which was not possible in this study. Further, the sample size for comparing performance against pRNFL is small, and confirmation in a larger and more diverse sample is necessary. Lastly, ON diagnosis was based on clinical records, which may have led to misclassification of some training data, if records were erroneous.

In summary we show that deep learning-based recognition of ON on peripapillary ring scans is feasible. While our study’s results are promising, further methodological improvements both regarding network architecture, used data modalities and training data variability are warranted.

## Data Availability

All data produced in the present study are available upon reasonable request to the authors.

## Acknowledgment

This study was supported in part by Berlin Institute of Health (project “DEEP-Neuroretina” to A.U. Brandt).

## Conflict of Interest

S. Motamedi received travel funding from Americas Committee for Treatment and Research in Multiple Sclerosis (ACTRIMS). S.K. Yadav is a cofounder and holds shares in technology start-up Nocturne GmbH, which has commercial interest in OCT applications in neurology; and is named as a coinventor on a pending patent application “Method and system for retinal tomography image quality”. J. Kauer-Bonin is currently an employee of Nocturne GmbH; and is named as a coinventor on a pending patent application “Method and system for retinal tomography image quality”. H.G. Zimmermann received research grants from Novartis and speaking honoraria from Bayer. S.L. Galetta received consulting fees from Biogen and Genentech. L.J. Balcer is the Editor-in-Chief of the Journal of Neuro-Ophthalmology; and is named as a coinventor on patent applications related to the Mobile Universal Lexicon Evaluation System (MULES) and the Staggered Uneven Number (SUN) tests owned by NYU, who also owns the copyright on these tests. F. Paul is a cofounder and holds shares in technology start-up Nocturne GmbH, which has commercial interest in OCT applications in neurology; served on the scientific advisory boards of Novartis and MedImmune; received travel funding and/or speaker honoraria from Bayer, Novartis, Biogen, Teva, Sanofi-Aventis/Genzyme, Merck Serono, Alexion, Chugai, MedImmune, and Shire; is an associate editor of Neurology: Neuroimmunology & Neuroinflammation; is an academic editor of PLoS ONE; consulted for Sanofi Genzyme, Biogen, MedImmune, Shire, and Alexion; received research support from Bayer, Novartis, Biogen, Teva, Sanofi-Aventis/Geynzme, Alexion, and Merck Serono; and received research support from the German Research Council, Werth Stiftung of the City of Cologne, German Ministry of Education and Research, Arthur Arnstein Stiftung Berlin, EU FP7 Framework Program, Arthur Arnstein Foundation Berlin, Guthy-Jackson Charitable Foundation, and NMSS. A.U. Brandt is a cofounder and holds shares in technology start-up Nocturne GmbH, which has commercial interest in OCT applications in neurology; is a cofounder and holds shares in Motognosis GmbH; received a grant from Berlin Institute of Health; received travel funding from Americas Committee for Treatment and Research in Multiple Sclerosis (ACTRIMS); named as a coinventor on several patent applications regarding retinal image analysis, MS serum biomarkers, MS-related myelination pathway, and motor function analysis; served on the observational study monitoring board of the CentrAl Vein Sign in MS (CAVS-MS) study (funded by the National Institutes of Health (NIH)); and serves as Secretary/Treasurer and Member of the Board of the International MS Visual System Consortium. All other authors have no conflicts of interest to declare.

## Notes

### Author Declarations

The study was approved by the local ethics committee at Charite - Universitaetsmedizin Berlin (EA1/182/10 and EA1/163/12). Independent validation data from NYU were collected under IRB approval at NYU. The study was conducted according to the Declaration of Helsinki in its currently applicable version and the applicable German and European laws. All participants gave written informed consent.

## References

1. Reich DS, Lucchinetti CF, Calabresi PA. Multiple sclerosis. 2018;378:169–180.

2. Poser CM, Paty DW, Scheinberg L, et al. New diagnostic criteria for multiple sclerosis: Guidelines for research protocols. Annals of Neurology 1983;13(3):227–231.

3. Thompson AJ, Banwell BL, Barkhof F, et al. Diagnosis of multiple sclerosis: 2017 revisions of the McDonald criteria. The Lancet Neurology 2018;17(2):162–173.

4. Van Der Vuurst De Vries RM, Mescheriakova JY, Wong YYM, et al. Application of the 2017 Revised McDonald Criteria for Multiple Sclerosis to Patients With a Typical Clinically Isolated Syndrome. JAMA Neurology 2018;75(11):1392.

5. Kaufmann M, Kuhle J, Puhan MA, et al. Factors associated with time from first-symptoms to diagnosis and treatment initiation of Multiple Sclerosis in Switzerland. Mult Scler J Exp Transl Clin 2018;4(4):2055217318814562.

6. Kaisey M, Solomon AJ, Luu M, et al. Incidence of multiple sclerosis misdiagnosis in referrals to two academic centers. Mult Scler Relat Disord 2019;30:51–56.

7. Kenney R, Liu M, Patil S, et al. Long-term outcomes in patients presenting with optic neuritis: Analyses of the MSBase registry. J Neurol Sci 2021;430:118067.

8. Toosy AT, Mason DF, Miller DH. Optic neuritis. The Lancet Neurology 2014;13(1):83–99.

9. Green AJ, McQuaid S, Hauser SL, et al. Ocular pathology in multiple sclerosis: retinal atrophy and inflammation irrespective of disease duration. Brain 2010;133(6):1591–1601.

10. Kerrison JB, Flynn T, Green WR. Retinal pathologic changes in multiple sclerosis. Retina 1994;14(5):445–51.

11. Ikuta F, Zimmerman HM. Distribution of plaques in seventy autopsy cases of multiple sclerosis in the United States. Neurology 1976;26(6 pt 2):26–8.

12. Toussaint D, Périer O, Verstappen A, Bervoets S. Clinicopathological study of the visual pathways, eyes, and cerebral hemispheres in 32 cases of disseminated sclerosis. Journal of Clinical Neuro-Ophthalmology 1983;3(3):211–20.

13. Brownlee WJ, Galetta S. Optic Nerve in Multiple Sclerosis Diagnostic Criteria: An Aye to the Eyes? Neurology 2021;96(4):139–140.

14. Filippi M, Preziosa P, Meani A, et al. Prediction of a multiple sclerosis diagnosis in patients with clinically isolated syndrome using the 2016 MAGNIMS and 2010 McDonald criteria: a retrospective study. The Lancet Neurology 2018;17(2):133–142.

15. Brandt AU, Martinez-Lapiscina EH, Nolan R, Saidha S. Monitoring the Course of MS With Optical Coherence Tomography. Curr Treat Options Neurol 2017;19(4):15.

16. Petzold A, de Boer JF, Schippling S, et al. Optical coherence tomography in multiple sclerosis: a systematic review and meta-analysis. The Lancet Neurology 2010;9(9):921– 932.

17. Petzold A, Balcer LJ, Calabresi PA, et al. Retinal layer segmentation in multiple sclerosis: a systematic review and meta-analysis. Lancet Neurol 2017;16(10):797–812.

18. Costello F, Coupland S, Hodge W, et al. Quantifying axonal loss after optic neuritis with optical coherence tomography. Annals of Neurology 2006;59(6):963–969.

19. Bock M, Brandt AU, Dörr J, et al. Patterns of retinal nerve fiber layer loss in multiple sclerosis patients with or without optic neuritis and glaucoma patients. Clin Neurol Neurosurg 2010;112(8):647–52.

20. Motamedi S, Gawlik K, Ayadi N, et al. Normative Data and Minimally Detectable Change for Inner Retinal Layer Thicknesses Using a Semi-automated OCT Image Segmentation Pipeline. Front Neurol 2019;10:1117.

21. LeCun Y, Bengio Y, Hinton G. Deep learning. Nature 2015;521(7553):436–444.

22. Ting DSW, Pasquale LR, Peng L, et al. Artificial intelligence and deep learning in ophthalmology. The British journal of ophthalmology 2019;103(2):167–175.

23. Kauer-Bonin J, Yadav SK, Beckers I, et al. Modular deep neural networks for automatic quality control of retinal optical coherence tomography scans. Comput Biol Med 2021;104822.

24. Yadav SK, Kafieh R, Zimmermann HG, et al. Deep Learning based Intraretinal Layer Segmentation using Cascaded Compressed U-Net. medRxiv 2021;2021.11.19.21266592.

25. Schippling S, Balk LJ, Costello F, et al. Quality control for retinal OCT in multiple sclerosis: validation of the OSCAR-IB criteria. Mult Scler 2015;21(2):163–170.

26. Aytulun A, Cruz-Herranz A, Aktas O, et al. The APO 2.0 Recommendations for Reporting Quantitative Optical Coherence Tomography Studies [Internet]. Neurology 2021;Available from: https://n.neurology.org/content/neurology/early/2021/04/28/WNL.0000000000012125.full.pdf

27. Petzold A, Albrecht P, Balcer L, et al. Artificial intelligence extension of the OSCAR-IB criteria. Annals of Clinical and Translational Neurology 2021;8(7):1528–1542.

28. R Core Team. R: A Language and Environment for Statistical Computing [Internet]. Vienna, Austria: R Foundation for Statistical Computing; 2018.Available from: https://www.R-project.org/

29. Yu F, Koltun V, Funkhouser T. Dilated residual networks. In: Proceedings of the IEEE conference on computer vision and pattern recognition. 2017 p. 472–480.

30. He K, Zhang X, Ren S, Sun J. Deep residual learning for image recognition. In: Proceedings of the IEEE conference on computer vision and pattern recognition. 2016 p. 770–778.

31. Nolan RC, Galetta SL, Frohman TC, et al. Optimal Intereye Difference Thresholds in Retinal Nerve Fiber Layer Thickness for Predicting a Unilateral Optic Nerve Lesion in Multiple Sclerosis. Journal of Neuro-Ophthalmology 2018;1–1.

32. Nolan-Kenney RC, Liu M, Akhand O, et al. Optimal intereye difference thresholds by optical coherence tomography in multiple sclerosis: An international study. Annals of Neurology 2019;85(5):618–629.

33. Coric D, Balk LJ, Uitdehaag BMJ, Petzold A. Diagnostic accuracy of optical coherence tomography inter-eye percentage difference for optic neuritis in multiple sclerosis. European Journal of Neurology 2017;24(12):1479–1484.

34. Petzold A, Chua SYL, Khawaja AP, et al. Retinal asymmetry in multiple sclerosis. Brain 2021;144(1):224–235.

35. Brandt AU, Specovius S, Oberwahrenbrock T, et al. Frequent retinal ganglion cell damage after acute optic neuritis. Mult Scler Relat Disord 2018;22:141–147.

36. Outteryck O, Lopes R, Drumez É, et al. Optical coherence tomography for detection of asymptomatic optic nerve lesions in clinically isolated syndrome. Neurology 2020;95(6):e733–e744.

37. Vidal-Jordana A, Rovira A, Arrambide G, et al. Optic Nerve Topography in Multiple Sclerosis Diagnosis. Neurology 2021;96(4):e482–e490.

